## Supplementary Materials for "Towards estimating true cholera burden: a systematic review and meta-analysis of *Vibrio cholerae* positivity"


**Supplementary Methods** **1**

Systematic review search terms 1

Statistical model 3

Estimation of *V. cholerae* positivity overall and by study methodology 3

**Supplementary Figures** **4**

Figure S1. Data coverage by geography 4

Figure S2. Data coverage over time 5

Figure S3. *Vibrio cholerae* positivity by incidence and suspected case characteristics 6

Figure S4. Posterior distributions of *V. cholerae* positivity 7

**Supplementary Tables** **8**

Table S1. Priors used in the latent class meta-analysis to estimate sensitivity and specificity of each diagnostic test 8

Table S2. Characteristics of suspected cholera cases reported in each observation 9

Table S3. Estimated sensitivity and specificity of each diagnostic test 9

Table S4. Estimated underlying *V. cholerae* positivity 10

Table S5. Odds of *V. cholerae* positivity by age and outbreak context 10

**References** **11**

### Supplementary Methods

#### Systematic review search terms

<u>Original search</u>
We searched PubMed, Embase, Scopus, Google Scholar and *medRxiv* on October 16, 2021 using the following search terms:

**PubMed (4034 results):**
(efficacy[Title/Abstract] OR effectiveness[Title/Abstract] OR surveillance[Title/Abstract] OR outbreak*[Title/Abstract] OR diagnos*[Title/Abstract] OR confirm*[Title/Abstract] OR incidence[Title/Abstract] OR PCR[Title/Abstract] OR "rapid diagnostic"[Title/Abstract] OR "rapid detection"[Title/Abstract] OR "rapid test*"[Title/Abstract] OR RDT[Title/Abstract] OR RDTs OR culture[Title/Abstract]) AND (cholera*[Title/Abstract] OR cholera[MeSH]) NOT ("animals"[MeSH Terms] NOT "humans"[MeSH Terms])

**Embase (4653 results):**
(effectiveness:ab,ti OR efficacy:ab,ti OR surveillance:ab,ti OR outbreak*:ab,ti OR diagnos*:ab,ti OR confirm*:ab,ti OR incidence:ab,ti OR pcr:ab,ti OR 'rapid diagnostic':ab,ti OR 'rapid detection':ab,ti OR 'rapid test*':ab,ti OR rdt:ab,ti OR rdts:ab,ti OR culture:ab,ti) AND cholera*:ab,ti AND [2000-2021]/py NOT ('animal'/exp NOT 'human'/exp)

**Scopus (2415 results):**
TITLE-ABS(effectiveness OR efficacy OR surveillance OR outbreak* OR diagnos* OR confirm* OR incidence OR pcr OR "rapid diagnostic" OR "rapid detection" OR "rapid test*" OR rdt OR culture ) AND TITLE-ABS (cholera*) AND PUBYEAR > 2000 AND NOT INDEX (embase)

**Google Scholar (2180 results exported):**
(cholera OR cholerae) AND (effectiveness OR efficacy OR surveillance OR outbreak OR outbreaks OR confirm OR confirmed OR confirmation OR diagnose OR diagnosed OR diagnosis OR incidence OR PCR OR culture OR "rapid test" OR "rapid tests" OR "rapid diagnostic" OR "rapid detection" OR RDT OR RDTs)

**medRxiv (13 results exported):**
Simply searched “cholera” and reviewed all the 287 results, 13 were exported to endnote for possible inclusion.

In total, 8209 records were identified in this search for upload to Covidence. Using Covidence, 271 duplicates were removed, with 7938 left for screening.

Updated search
We updated the search on April 19, 2023 to include additional studies that had been published or indexed since our original search. We used the same search terms as above with the added restriction that data were entered into the database (PubMed and Embase) or published after October 16, 2021 (Scopus and *medRxiv*) [1]. These additions are highlighted in red below. We did not update the search for Google Scholar because we were not able to restrict the search by date studies were entered or indexed in the database.

**PubMed (417 results):**
(efficacy[Title/Abstract] OR effectiveness[Title/Abstract] OR surveillance[Title/Abstract] OR outbreak*[Title/Abstract] OR diagnos*[Title/Abstract] OR confirm*[Title/Abstract] OR incidence[Title/Abstract] OR PCR[Title/Abstract] OR "rapid diagnostic"[Title/Abstract] OR "rapid detection"[Title/Abstract] OR "rapid test*"[Title/Abstract] OR RDT[Title/Abstract] OR RDTs OR culture[Title/Abstract]) AND (cholera*[Title/Abstract] OR cholera[MeSH]) NOT ("animals"[MeSH Terms] NOT "humans"[MeSH Terms]) AND ("2021/10/16"[EDAT] : "3000"[EDAT])

**Embase (246 results):**
(effectiveness:ab,ti OR efficacy:ab,ti OR surveillance:ab,ti OR outbreak*:ab,ti OR diagnos*:ab,ti OR confirm*:ab,ti OR incidence:ab,ti OR pcr:ab,ti OR 'rapid diagnostic':ab,ti OR 'rapid detection':ab,ti OR 'rapid test*':ab,ti OR rdt:ab,ti OR rdts:ab,ti OR culture:ab,ti) AND cholera*:ab,ti AND [2000-2021]/py NOT ('animal'/exp NOT 'human'/exp) AND [16-10-2021]/sd NOT [20-04-2023]/sd

**Scopus (277 results):**
TITLE-ABS(effectiveness OR efficacy OR surveillance OR outbreak* OR diagnos* OR confirm* OR incidence OR pcr OR "rapid diagnostic" OR "rapid detection" OR "rapid test*" OR rdt OR culture ) AND TITLE-ABS (cholera*) AND ( PUBYEAR > 2021 OR PUBDATETXT ( october 2021 ) OR PUBDATETXT ( november 2021 ) OR PUBDATETXT ( december 2021 ) ) AND NOT INDEX (embase)

***medRxiv* (4 results exported):**
Searched “cholera” and restricted the data posting date after Oct-16-2021 and reviewed all the 146 results, 4 were exported to endnote for possible inclusion.

In total, 944 new records identified in this updated search. After uploaded to Covidence, 157 duplicates were removed, with 787 left for screening.

Note on *medRxiv* and pre-prints
Although we originally included pre-prints in our screening, we excluded pre-prints that had not been peer-reviewed by the time of the updated search in our final analyses or that no longer had cholera positivity data in the published version of the manuscript (as was the case in the one pre-print study that was initially included).

#### Statistical model

To estimate proportion of suspected cases that are true *V. cholerae* infections, we performed a hierarchical meta-analysis using CmdStanR version 0.5.2 as an interface to Stan for R [2,3].

In this model, $p_{obs}(i,j)$, probability of observing a positive test result by test $j$ for observation $i$ is a function of the true percent positive $p_{pos}(i)$ of observation $i$ and test sensitivity $\theta^{+}$ and specificity $\theta^{-}$ of test $j$:

$$p_{obs}(i,j) = p_{pos}(i)\ \theta^{+}(j)\ +\ (1 - p_{pos}(i))\ (1 - \theta^{-}(j))$$
$$n_{obs}(i,j) \sim Binomial(N_{obs}(i,j), p_{obs}(i,j)),$$

where $N_{obs}(i,j)$ is the number of suspected cases tested and $n_{obs}(i,j)$ is the number that tested positive. The true percent positive $p_{pos}$ for each observation was modeled as a function of covariates $X$ (i.e. sampling strategy for test, age constraint in case definition, whether or not surveillance was initiated in response to an outbreak) and a random effect by observation, $\varepsilon(i)$ :

$$logit\left(p_{pos}(i)\right) = \alpha + X(i)\beta\ + \varepsilon(i)$$

We used a $Normal(0,2)$ prior on the coefficients $\beta$ and the global intercept $\alpha$ and a standard normal distribution prior on the random effects. In sensitivity analysis, we used a $Normal(0.9,2)$ prior the global intercept $\alpha$.

In each iteration of this model, we used a randomly selected draw from the posterior distribution of sensitivity and specificity in the JAGS model output (described in main text) to incorporate uncertainty in test performance. We ran 3,000 total iterations for all models implemented with Stan, including 4 chains each with 2,000 sampling iterations and 1,000 warm-up iterations. Convergence of all models was assessed by R-hat values and visual inspection of traceplots.

#### Estimation of *V. cholerae* positivity overall and by study methodology

To estimate *V. cholerae* positivity across all studies $p$, we estimated the proportion of suspected cases that represented true *V. cholerae* infections we marginalized over the study-level random effects for specific strata, s, following similar methods to [4] as follows:

$$p_s = \int_0^1 logit^{-1}(\ \alpha + X_s\beta + \sigma * \Phi^{-1}(t))dt,$$

Where, $\Phi^{-1}(t)$ is the normal distribution quantile function and $\sigma$ is the variance of the random effect distribution.

#### Supplementary Figures

##### Figure S1. Data coverage by geography

Number of observations in the primary dataset at each administrative level as defined by GADM (https://gadm.org/) by country. Countries with >10 observations displayed as 10. Haiti includes 1 observation at the national level, 2 at the first administrative division, and 4 at the second.

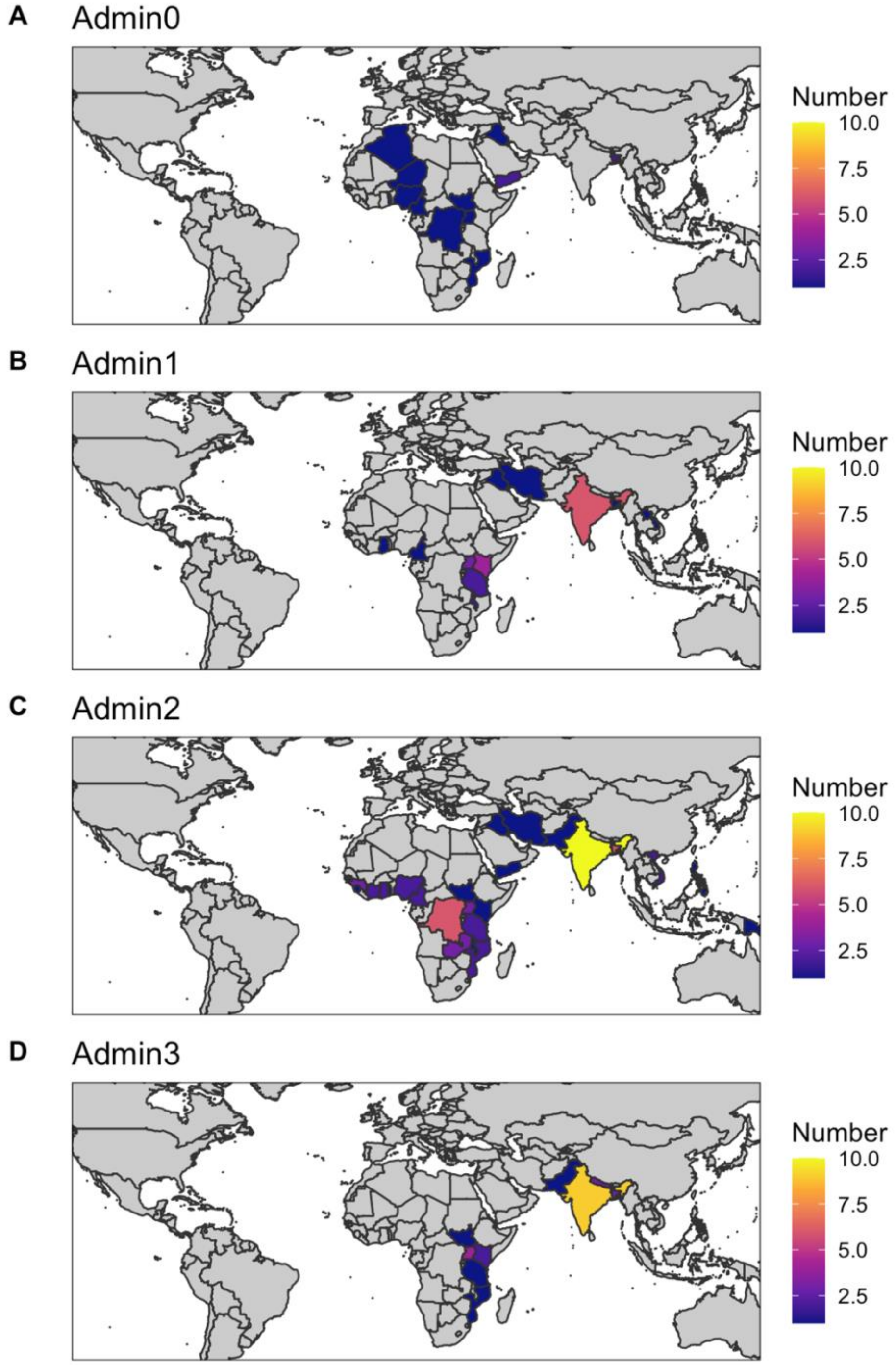

#### Figure S2. Data coverage over time

Number of observations in the primary dataset at each administrative level as defined by GADM (https://gadm.org/) by country within different time periods. Countries with >10 observations displayed as 10. Year represents the year sampling was completed. Excludes 9 studies missing a study end date. Haiti includes 3 studies during 2010-2014 and 4 during 2015-2022.

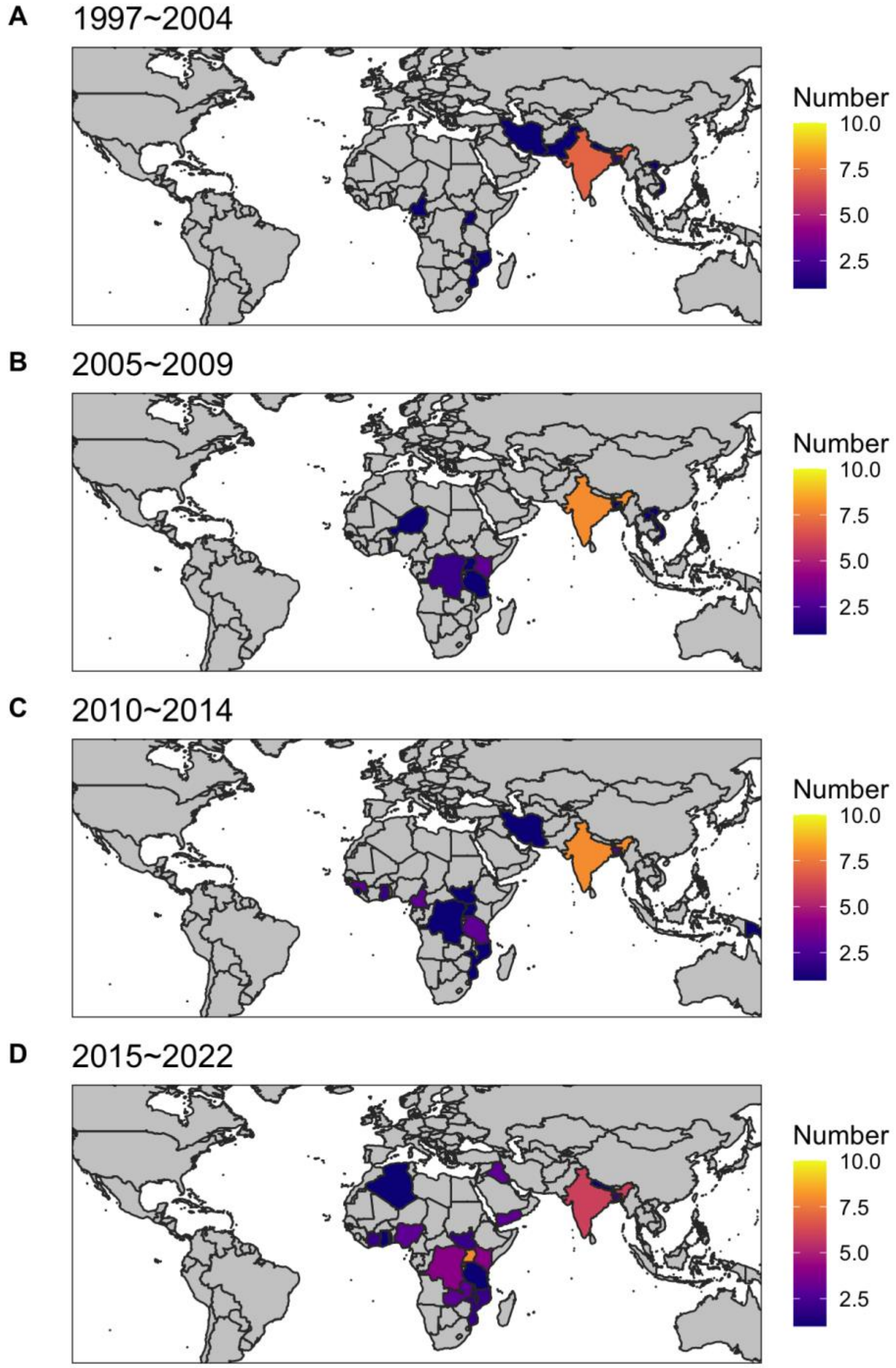

#### Figure S3. *Vibrio cholerae* positivity by incidence and suspected case characteristics

Relationship between reported *V. cholerae* positivity and **A)** proportion of suspected cholera cases tests that were under 5 years of age, **B)** suspected cholera incidence rate per 10,000 at each study site in Africa, **C)** proportion of suspected cases severely dehydrated, and **D)** proportion of suspected cases on antibiotics prior to testing. Size of the points is proportional to the number of cases tested, and shapes indicate which diagnostic test was used to confirm *V. cholerae* infection. Confidence intervals for Spearman rank correlation coefficients estimated using bootstrapping (nrep=1000). Smoothing method is loess (without weights). In **B)** estimated suspected cholera incidence rates in Africa for 2010-2016 [5] were aggregated to the administrative division that best represented each study's catchment area by dividing the total estimated cholera cases in each area by its estimated population.

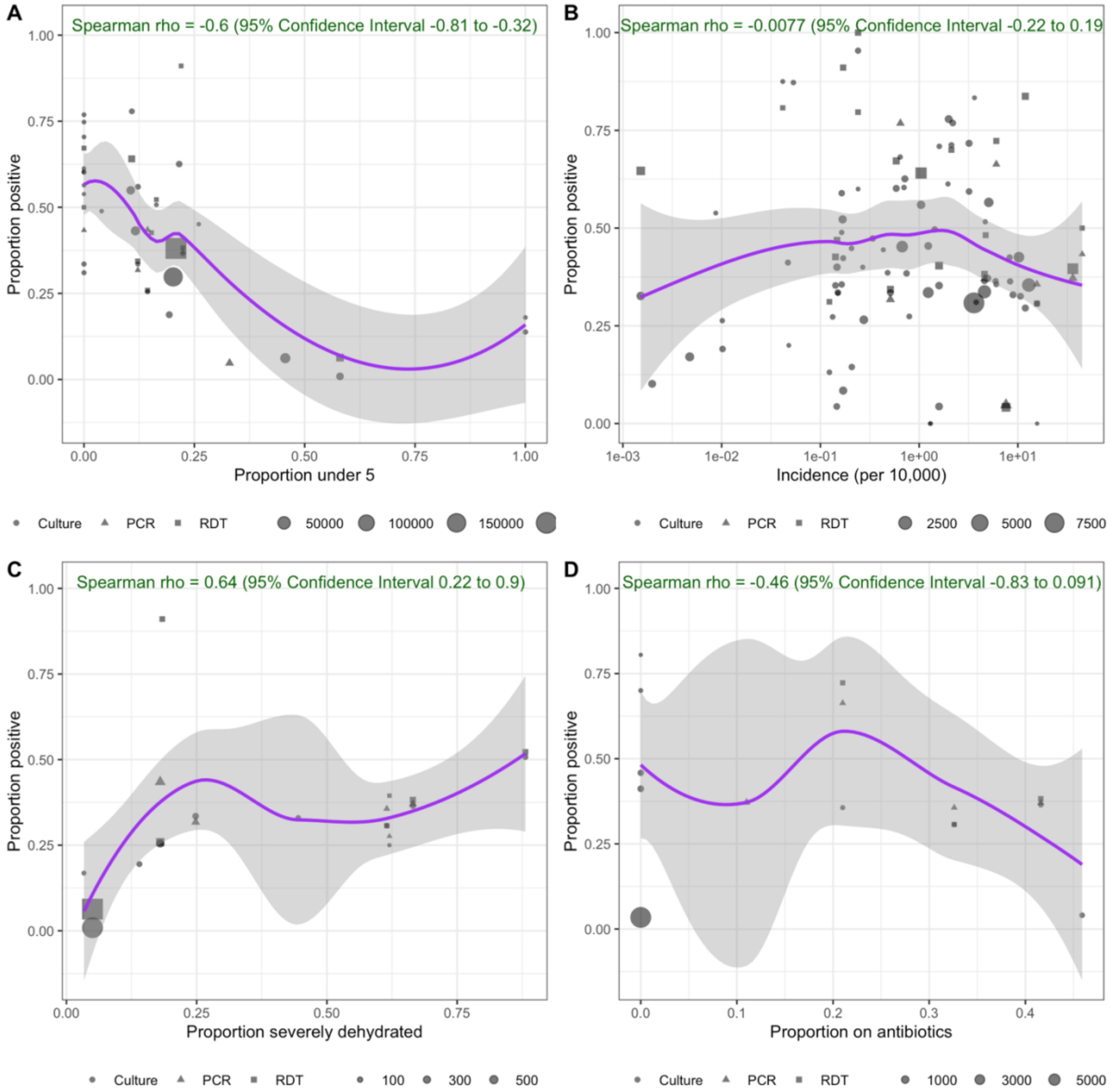

#### Figure S4. Posterior distributions of *V. cholerae* positivity

Posterior distributions of *V. cholerae* positivity estimated using the random-effects model corresponding to results in Table S4. **A)** Unadjusted. **B)** Adjusted for test performance. **C)** Adjusted for test performance, sensitivity analysis shifting prior on alpha from Normal(0,2) as in A-B to Normal(0.9,2). On the left, prior (green) and posterior distribution (purple) of the global intercept are shown in logit space. On the right, histograms of estimated *V. cholerae* positivity.

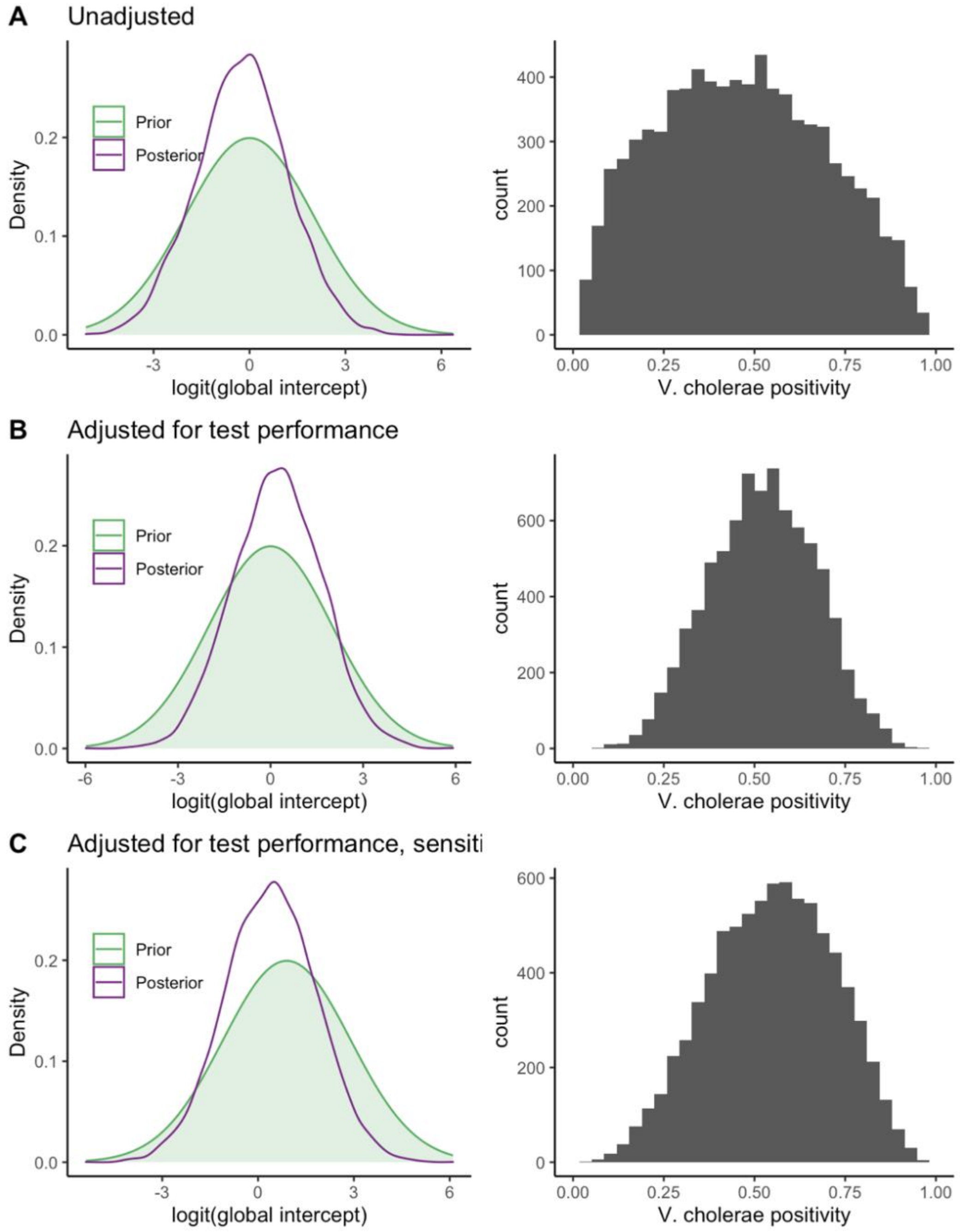

#### Supplementary Tables

Table S1. Priors used in the latent class meta-analysis to estimate sensitivity and specificity of each diagnostic test

| | Prior distribution in JAGS model | Lower bound of truncation | Upper bound of truncation |
|---|---|---|---|
| Latent prevalence of cholera infection π | $\pi \sim beta(1,1)$ | none | none |
| Sensitivity of culture $\mu_{sen(culture)}$ | $\mu_{sen(culture)} \sim beta(1,1)$ | 0.3 | 1 |
| Sensitivity of PCR† $\mu_{sen(PCR)}$ | $\mu_{sen(PCR)} \sim beta(1,1)$ | 0.5 | 1 |
| Sensitivity of RDT* $\mu_{sen(RDT)}$ | $\mu_{sen(RDT)} \sim beta(1,1)$ | 0.5 | 1 |
| Specificity of culture $\mu_{spec(culture)}$ | $\mu_{spec(culture)} \sim beta(1,1)$ | 0.8 | 1 |
| Specificity of PCR† $\mu_{spec(PCR)}$ | $\mu_{spec(PCR)} \sim beta(1,1)$ | 0.8 | 1 |
| Specificity of RDT* $\mu_{spec(RDT)}$ | $\mu_{spec(RDT)} \sim beta(1,1)$ | 0.5 | 1 |

†PCR = Polymerase Chain Reaction

*RDT = Rapid Diagnostic Test

Table S2. Characteristics of suspected cholera cases reported in each observation

| Characteristic | Reported | Total observations | Percent |
|---|---|---|---|
| Proportion vaccinated | No | 127 | 96.2 |
| | Yes | 5 | 3.8 |
| Proportion under 5 years old | No | 100 | 75.8 |
| | Yes | 32 | 24.2 |
| Proportion under 15 years old | No | 114 | 86.4 |
| | Yes | 18 | 13.6 |
| Proportion severely dehydrated | No | 121 | 91.7 |
| | Yes | 11 | 8.3 |
| Proportion moderately dehydrated | No | 123 | 93.2 |
| | Yes | 9 | 6.8 |
| Proportion female | No | 104 | 78.8 |
| | Yes | 28 | 21.2 |
| Proportion that received antibiotics prior to confirmation test | No | 122 | 92.4 |
| | Yes | 10 | 7.6 |
| Proportion under 5 years old & proportion severely dehydrated | No | 126 | 95.5 |
| | Yes | 6 | 4.5 |
| Proportion under 5 years old, severely dehydrated, & received antibiotics prior to test | No | 131 | 99.2 |
| | Yes | 1 | 0.8 |

Table S3. Estimated sensitivity and specificity of each diagnostic test

Estimate is median sensitivity and specificity pooled across four studies that reported results for all diagnostics tests, as described in Methods. Parentheses show 95% Credible Interval.

| Measure | Test | Estimate (%) |
|---|---|---|
| Sensitivity | Culture | 82.0 (37.5, 98.7) |
| | PCR† | 85.1 (53.6, 98.9) |
| | RDT* | 90.4 (55.2, 99.5) |
| Specificity | Culture | 94.3 (81.5, 99.6) |
| | PCR† | 94.2 (81.1, 99.7) |
| | RDT* | 88.9 (54.8, 99.4) |

†PCR = Polymerase Chain Reaction

*RDT = Rapid Diagnostic Test

#### Table S4. Estimated underlying *V. cholerae* positivity

"Unadjusted" is mean *V. cholerae* positivity (95% credible interval) from random effects meta-analysis without adjustments for test performance. "Adjusted" refers to *V. cholerae* estimates additionally adjusted for test performance, where the primary analysis includes a Normal(0,2) prior on the global intercept and a sensitivity analysis includes the prior shifted to Normal(0.9,2). "Stratified estimate for high quality…" corresponds to post-stratified estimates of *V. cholerae* positivity for studies that use high quality sampling methods and whether an age minimum was set in the suspected case definition, as well as whether surveillance was initiated in response to an outbreak.

| Setting | Version | Positivity (%) |
|---|---|---|
| Across all settings | Unadjusted | 46.4 (7.7, 89.2) |
| | Adjusted for test performance | 52.2 (24.2, 79.8) |
| | Adjusted for test performance, prior on alpha shifted (sensitivity analysis) | 54.8 (21, 85.3) |
| High quality, stratified by inclusion of age restrictions | Stratified estimate for high quality sample with no known age restriction | 45.9 (18.8, 75.5) |
| | Stratified estimate for high quality sample with any age restriction | 68.2 (32.6, 97.6) |
| High quality, stratified by whether surveillance was initiated in response to outbreak | Stratified estimate for high quality sample during non-outbreak surveillance | 41.7 (11.9, 77) |
| | Stratified estimate for high quality sample during outbreak surveillance | 78.2 (39.7, 99.1) |

#### Table S5. Odds of *V. cholerae* positivity by age and outbreak context

Odds that a suspected cholera case seeking testing or care has a true *V. cholerae* O1/O139 infection with low sampling quality compared to high sampling quality, with any minimum age was set in case definition, and with surveillance initiated in response to an outbreak compared to non-outbreak surveillance (i.e., routine or post-vaccination surveillance).

| Variable | Odds ratio |
|---|---|
| Low sampling quality | 3.80 (0.95, 9.86) |
| Minimum age in case definition | 2.33 (0.54, 6.40) |
| Outbreak surveillance | 5.71 (1.53, 15.43) |
