## Supplementary material for "Towards estimating true cholera burden: a systematic review and meta-analysis of *Vibrio cholerae* positivity": S1 Checklist

### PRISMA 2020 Checklist

| Section and Topic | Item # | Checklist item | Location where item is reported |
|---|---|---|---|
| **TITLE** | | | |
| Title | 1 | Identify the report as a systematic review. | **Title** |
| **ABSTRACT** | | | |
| Abstract | 2 | See the PRISMA 2020 for Abstracts checklist. | **Abstract** |
| **INTRODUCTION** | | | |
| Rationale | 3 | Describe the rationale for the review in the context of existing knowledge. | **Introduction**, paragraphs 1-2 |
| Objectives | 4 | Provide an explicit statement of the objective(s) or question(s) the review addresses. | **Introduction**, paragraph 3 |
| **METHODS** | | | |
| Eligibility criteria | 5 | Specify the inclusion and exclusion criteria for the review and how studies were grouped for the syntheses. | **Methods**, section "Systematic review", paragraph 1 |
| Information sources | 6 | Specify all databases, registers, websites, organisations, reference lists and other sources searched or consulted to identify studies. Specify the date when each source was last searched or consulted. | **Supplementary methods**, section "Systematic review search terms" |
| Search strategy | 7 | Present the full search strategies for all databases, registers and websites, including any filters and limits used. | **Supplementary methods**, section "Systematic review search terms" |
| Selection process | 8 | Specify the methods used to decide whether a study met the inclusion criteria of the review, including how many reviewers screened each record and each report retrieved, whether they worked independently, and if applicable, details of automation tools used in the process. | **Methods**, section "Systematic review", paragraph 2 |
| Data collection process | 9 | Specify the methods used to collect data from reports, including how many reviewers collected data from each report, whether they worked independently, any processes for obtaining or confirming data from study investigators, and if applicable, details of automation tools used in the process. | **Methods**, section "Systematic review", paragraph 2 |
| Data items | 10a | List and define all outcomes for which data were sought. Specify whether all results that were compatible with each outcome domain in each study were sought (e.g. for all measures, time points, analyses), and if not, the methods used to decide which results to collect. | **Methods**, section "Systematic review", paragraph 2 |

### PRISMA 2020 Checklist

| Section and Topic | Item # | Checklist item | Location where item is reported |
| --- | --- | --- | --- |
| | | | and **Supplementary Data 1** |
| | 10b | List and define all other variables for which data were sought (e.g. participant and intervention characteristics, funding sources). Describe any assumptions made about any missing or unclear information. | **Methods**, section "Systematic review", paragraphs 2-3 and **Supplementary Data 1** |
| Study risk of bias assessment | 11 | Specify the methods used to assess risk of bias in the included studies, including details of the tool(s) used, how many reviewers assessed each study and whether they worked independently, and if applicable, details of automation tools used in the process. | **Methods,** section "Systematic review", paragraph 5 |
| Effect measures | 12 | Specify for each outcome the effect measure(s) (e.g. risk ratio, mean difference) used in the synthesis or presentation of results. | **Methods**, section "Terminology" and section "Data analysis", sub-section *Estimating* V. cholerae *positivity and sources of heterogeneity*, paragraph 1 |
| Synthesis methods | 13a | Describe the processes used to decide which studies were eligible for each synthesis (e.g. tabulating the study intervention characteristics and comparing against the planned groups for each synthesis (item #5)). | These were the same as the inclusion criteria: **Methods**, section "Systematic review", paragraphs 1-3 |
| | 13b | Describe any methods required to prepare the data for presentation or synthesis, such as handling of missing summary statistics, or data conversions. | **Methods**, section "Systematic review", paragraph 2 |
| | 13c | Describe any methods used to tabulate or visually display results of individual studies and syntheses. | **Methods**, section |

### PRISMA 2020 Checklist

| Section and Topic | Item # | Checklist item | Location where item is reported |
|---|---|---|---|
| | | | "Systematic review", paragraph 5 |
| | 13d | Describe any methods used to synthesize results and provide a rationale for the choice(s). If meta-analysis was performed, describe the model(s), method(s) to identify the presence and extent of statistical heterogeneity, and software package(s) used. | **Methods** section, "Data analysis", sub-section *Estimating* V. cholerae *positivity and sources of heterogeneity*, paragraph 1 |
| | 13e | Describe any methods used to explore possible causes of heterogeneity among study results (e.g. subgroup analysis, meta-regression). | **Methods** section, "Data analysis", sub-section *Estimating* V. cholerae *positivity and sources of heterogeneity*, paragraphs 1-2 |
| | 13f | Describe any sensitivity analyses conducted to assess robustness of the synthesized results. | **Methods**, section "Data analysis", sub-section *Estimating* V. cholerae *positivity and sources of heterogeneity*, paragraph 1 |
| Reporting bias assessment | 14 | Describe any methods used to assess risk of bias due to missing results in a synthesis (arising from reporting biases). | Not applicable (missing data were generally not reported by the included studies) |
| Certainty assessment | 15 | Describe any methods used to assess certainty (or confidence) in the body of evidence for an outcome. | We did not conduct a certainty assessment |
| **RESULTS** | | | |

### PRISMA 2020 Checklist

| Section and Topic | Item # | Checklist item | Location where item is reported |
|---|---|---|---|
| Study selection | 16a | Describe the results of the search and selection process, from the number of records identified in the search to the number of studies included in the review, ideally using a flow diagram. | **Results**, paragraph 1 and Figure 1 |
| | 16b | Cite studies that might appear to meet the inclusion criteria, but which were excluded, and explain why they were excluded. | **Results**, paragraph 1, References |
| Study characteristics | 17 | Cite each included study and present its characteristics. | **Results**, paragraph 1, References, Supplementary data 1, Table 1 |
| Risk of bias in studies | 18 | Present assessments of risk of bias for each included study. | We did not formally assess risk of bias for each of the individual 131 studies initially identified for inclusion |
| Results of individual studies | 19 | For all outcomes, present, for each study: (a) summary statistics for each group (where appropriate) and (b) an effect estimate and its precision (e.g. confidence/credible interval), ideally using structured tables or plots. | Figure 3, Figure 4, and Table S4 |
| Results of syntheses | 20a | For each synthesis, briefly summarise the characteristics and risk of bias among contributing studies. | **Results**, paragraphs 1-3, and Figures S1-S2 and Table S2 |
| | 20b | Present results of all statistical syntheses conducted. If meta-analysis was done, present for each the summary estimate and its precision (e.g. confidence/credible interval) and measures of statistical heterogeneity. If comparing groups, describe the direction of the effect. | **Results**, section "Adjusted underlying *V. cholerae* positivity", paragraphs 2-3, and Figure 3, Figure S4, and Table S4 |
| | 20c | Present results of all investigations of possible causes of heterogeneity among study results. | **Results**, section "Factors associated with variation in *V. cholerae* positivity", paragraphs 1-2, |

### PRISMA 2020 Checklist

| Section and Topic | Item # | Checklist item | Location where item is reported |
|---|---|---|---|
| | | | and Tables S5-S6 |
| | 20d | Present results of all sensitivity analyses conducted to assess the robustness of the synthesized results. | **Results**, section "<u>Factors associated with variation in *V. cholerae* positivity</u>", paragraphs 1-2, and Table S4 |
| Reporting biases | 21 | Present assessments of risk of bias due to missing results (arising from reporting biases) for each synthesis assessed. | Not applicable |
| Certainty of evidence | 22 | Present assessments of certainty (or confidence) in the body of evidence for each outcome assessed. | A formal certainty assessment was not conducted. "Confidence" is represented by 95% Credible Intervals (95% CI) throughout the abstract, results, figures, and tables. |
| **DISCUSSION** | | | |
| Discussion | 23a | Provide a general interpretation of the results in the context of other evidence. | **Discussion**, paragraphs 1-2 |
| | 23b | Discuss any limitations of the evidence included in the review. | **Discussion**, paragraph 3 |
| | 23c | Discuss any limitations of the review processes used. | **Discussion**, paragraph 3 |
| | 23d | Discuss implications of the results for practice, policy, and future research. | **Discussion**, paragraphs 4-5 |
| **OTHER INFORMATION** | | | |
| Registration and protocol | 24a | Provide registration information for the review, including register name and registration number, or state that the review was not registered. | **Methods**, section "<u>Systematic review</u>", paragraph 4 |
| | 24b | Indicate where the review protocol can be accessed, or state that a protocol was not prepared. | **Methods**, section "<u>Systematic</u> |

| Section and Topic | Item # | Checklist item | Location where item is reported |
|---|---|---|---|
| | | | review”, paragraph 4 |
| | 24c | Describe and explain any amendments to information provided at registration or in the protocol. | Not applicable |
| Support | 25 | Describe sources of financial or non-financial support for the review, and the role of the funders or sponsors in the review. | **Funding** section |
| Competing interests | 26 | Declare any competing interests of review authors. | **Conflicts of Interest** section |
| Availability of data, code and other materials | 27 | Report which of the following are publicly available and where they can be found: template data collection forms; data extracted from included studies; data used for all analyses; analytic code; any other materials used in the review. | **Methods**, section “Data analysis”, sub-section *Data availability*, and **Supplementary Data 1** |
